# Sociodemographic factors affecting not receiving COVID-19 vaccine in Japan among people who originally intended to vaccinate: a prospective cohort study

**DOI:** 10.1101/2022.07.29.22277984

**Authors:** Akiko Matsuyama, Takahiro Mori, Akira Ogami, Kosuke Mafune, Seiichiro Tateishi, Mami Kuwamura, Keiji Muramatsu, Yoshihisa Fujino, Koji Mori, the CORoNaWork project

## Abstract

**Background:** Vaccine hesitancy is a major issue for acquiring herd immunity. However, some individuals may go unvaccinated owing to inhibitory factors other than vaccine hesitancy. If there is even a small number of such people, support is needed for equitable vaccine distribution and acquiring herd immunity. We investigated sociodemographic factors that affected not undergoing COVID-19 vaccination in Japan among individuals who initially had strong intention to vaccinate.

**Methods:** We conducted this prospective cohort study on workers aged 20–65 years from December 2020 (baseline), to December 2021 using a self-administered questionnaire survey. There were 27,036 participants at baseline and 18,560 at follow-up. We included 6,955 participants who answered yes to this question at baseline: “Would you like to receive a COVID-19 vaccine as soon as it becomes available?” We applied multilevel logistic regression analyses to examine the association between sociodemographic factors and being unvaccinated at follow-up.

**Results:** In all, 289 participants (4.2%) went unvaccinated. The odds ratios (ORs) for being unvaccinated were significantly higher for participants aged 30–39 and 40–49 than those aged 60–65 years. Being divorced, widowed, or single, having low income, and having COVID-19 infection experience also had higher ORs.

**Conclusions:** We found that some participants who initially had strong intention to vaccinate may have gone unvaccinated owing to vaccine side effects and the financial impact of absenteeism due to side effects. It is necessary to provide information repeatedly about the need for vaccination as well as social support to ensure that those who intend to vaccinate are able to do so.

## 1. Introduction

As with many viral infections, vaccination against COVID-19 is the most effective infection control[1]. Various types of vaccines for COVID-19, including mRNA vaccines, have been developed over a short period of time and applied worldwide since December 2020 [2], [3]. The efficacy and side effects differ according to the type of vaccine; however, many vaccines have been shown both to prevent onset and avert severity and death [4], [5], [6].

With an emerging epidemic like COVID-19, a high inoculation rate of effective vaccines can be used to establish herd immunity and control it [7]. However, vaccine hesitancy—defined as a “delay in acceptance or refusal of vaccination despite availability of vaccination services”—hinders effective vaccination and poses a major public health issue [8]. To reduce such hesitancy for COVID-19 in Japan, educational activities about the significance and risks of vaccination, free vaccination, and fair vaccine distribution began on February 17, 2021 for health-care workers; they were followed by older people and individuals with underlying medical conditions [9]. Also, efforts have been made to create convenient vaccination sites and provide vaccines in the workplace [3]. By December 2021, the second-dose vaccination among people aged over 12 years had been mostly completed, amounting to 73.4% of the total population [10].

Even though some people initially intended to be vaccinated, they later refuse vaccination owing to some inhibitory factors such as income factors —even though they too initially intended to receive it. If there is even a small number of such people, support is needed for equitable vaccine distribution and acquiring herd immunity. There is a necessity for a prospective cohort study or trajectory study to confirm whether people who initially intended to be vaccinated actually did so; however, few studies have examined the factors that influence vaccination decision. Sigerl et al. investigated COVID-19 vaccination in the United States; they found that 7% of people who were willing to be vaccinated in the baseline survey had become unwilling by the follow-up survey; however, the authors did not confirm actual vaccination rates and did not investigate the related factors [11].

As part of the Collaborative Online Research on Novel-coronavirus and Work (CORoNaWork) study [12], we asked participants about their intention to receive COVID-19 vaccination at baseline and their actual vaccination at follow-up. Using those data, we conducted a trajectory study to examine the sociodemographic factors that affected the actual vaccination rate of individuals who at baseline expressed strong intention to undergo vaccination.

## 2. Methods

### 2.1 Study design

We undertook this prospective cohort study as part of the CORoNaWork study. We conducted the baseline survey on December 22–25, 2020 and the follow-up survey on December 15–22, 2021, by which time the second-dose vaccination had been almost completed in Japan [10]. The surveys were in the form of a self-administrated questionnaire using the Internet. Details of the study protocol are reported elsewhere [12]. The target participants were aged 20–65 years and employed at the time of the baseline survey (n = 33,087). Respondents were sampled taking into account region, occupation, and sex. After excluding 6,051 initial subjects who provided invalid responses, we finally included 27,036 in the database. The criteria for invalid responses were as follows: very short response time (≤6 minutes); extremely low body weight (<30 kg); extremely short height (<140 cm); inconsistent answers to similar questions throughout the survey; and incorrect answers to a question used solely to identify unreliable responses.

In all, 18,560 participants (68.6%) undertook the follow-up survey. We conducted our analysis on 6,955 participants who answered yes to this question in the baseline survey: “Would you like to receive a COVID-19 vaccine as soon as it becomes available?” Figure 1 is a flow diagram for this study.

**Figure 1.**
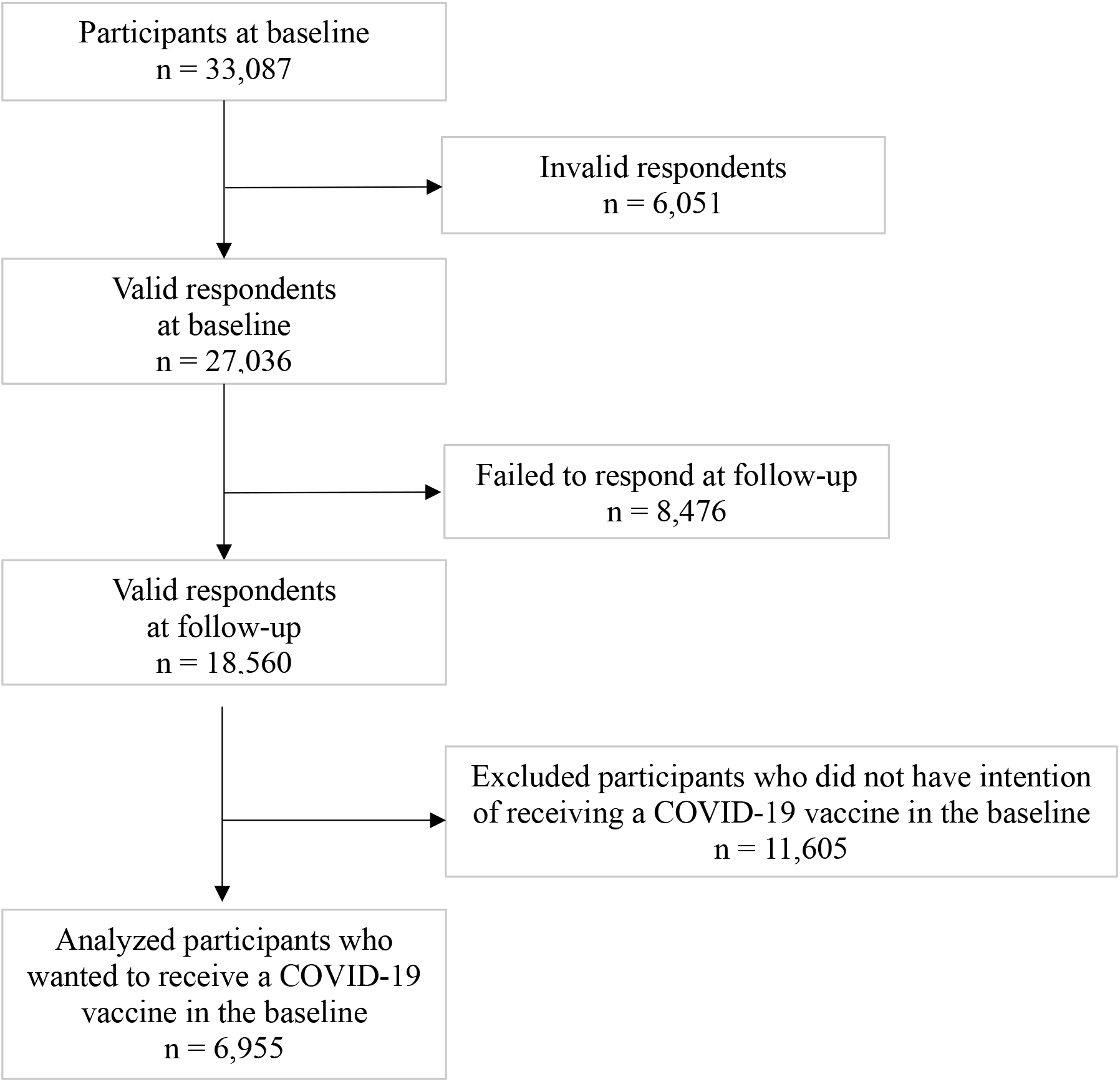
Flow diagram for this study

This study was approved by the Ethics Committee of the University of Occupational and Environmental Health, Japan (approval numbers: R2-079 and R3-006). Informed consent was obtained on the website from all participants.

### 2.2 COVID-19 vaccination status

We asked participants this question in the follow-up survey: “What is your COVID-19 vaccination status?” Participants chose one of three options: vaccinated twice; vaccinated once; and unvaccinated. We created a binary variable by defining “unvaccinated” as not having received a COVID-19 vaccine and the other options as having the vaccine.

### 2.3 Sociodemographic status

We investigated sex, age, marital status, annual household income, job type, and experience of COVID-19 infection. Age was classified into five groups: 20–29, 30–39, 40–49, 50–59, and 60–65 years. Marital status was classified into three categories: married, divorced or widowed, and single. Annual household income was classified into four groups: <4 million, 4–5.99 million, 6–8.99 million, and ≥9 million yen (US$1 equaled 109.75 yen in 2021) [13]. Job type was classified into three categories: mainly desk work, jobs mainly involving interpersonal communication, and mainly physical work. For experience of COVID-19 infection, we asked this question at baseline: “Have you ever been infected with COVID-19?” Respondents answered yes or no. At follow-up, we asked this question: “Have you been diagnosed with COVID-19 since January 2021?” Likewise, respondents answered yes or no. We defined participants who answered yes to either question as having experienced COVID-19.

### 2.4 Statistical analyses

We examined the association between sociodemographic factors and not receiving COVID-19 vaccination among respondents who had originally intended to do so. We estimated age-sex-adjusted odds ratios (ORs) and multivariate-adjusted ORs using a multilevel logistic regression model nested in the prefecture of residence to consider regional differences in the infection status of COVID-19. The multivariate model was adjusted for sex, age, marital status, annual household income, job type, and experience of COVID-19 infection. We did not adjust for education because doing so would have been an over-adjustment. We also conducted a trend test with age and annual household income as continuous variables. A *P* value of less than 0.05 was considered statistically significant. We conducted all analyses using Stata statistical software (release 16; StataCorp LLC, College Station, TX, USA).

## 3. Results

The characteristics of the participants appear in Table 1. Of the 6,955 participants analyzed, 4,382 were men (63.0%) and 2,573 women (37.0%). As of December 2021, 289 (4.2%) were unvaccinated.

**Table 1.**
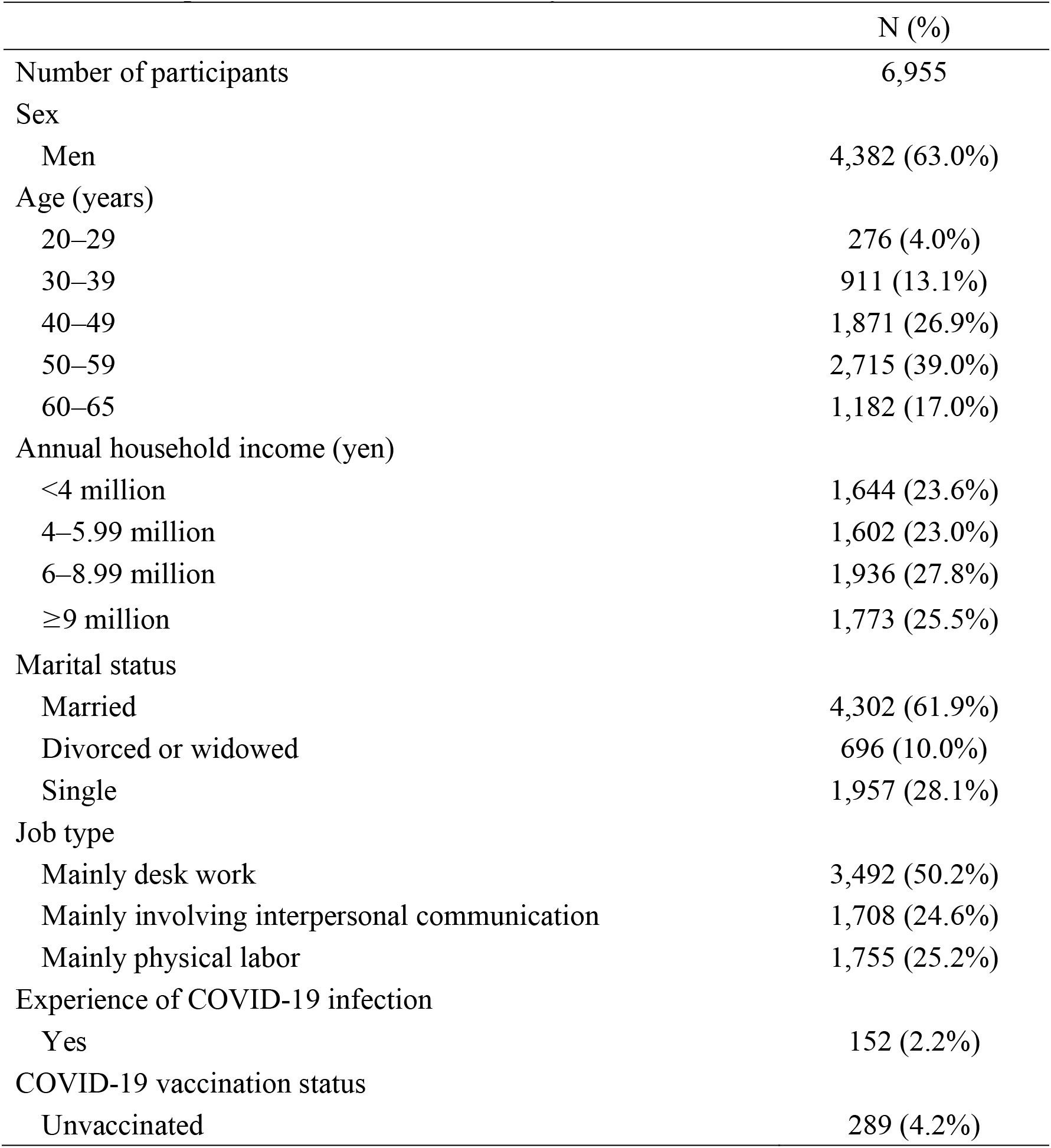
Participant characteristics of this study

|  | N (%) |
| --- | --- |
| Number of participants | 6,955 |
| Sex |  |
| Men | 4,382 (63.0%) |
| Age (years) |  |
| 20–29 | 276 (4.0%) |
| 30–39 | 911 (13.1%) |
| 40–49 | 1,871 (26.9%) |
| 50–59 | 2,715 (39.0%) |
| 60–65 | 1,182 (17.0%) |
| Annual household income (yen) |  |
| <4 million | 1,644 (23.6%) |
| 4–5.99 million | 1,602 (23.0%) |
| 6–8.99 million | 1,936 (27.8%) |
| ≥9 million | 1,773 (25.5%) |
| Marital status |  |
| Married | 4,302 (61.9%) |
| Divorced or widowed | 696 (10.0%) |
| Single | 1,957 (28.1%) |
| Job type |  |
| Mainly desk work | 3,492 (50.2%) |
| Mainly involving interpersonal communication | 1,708 (24.6%) |
| Mainly physical labor | 1,755 (25.2%) |
| Experience of COVID-19 infection |  |
| Yes | 152 (2.2%) |
| COVID-19 vaccination status |  |
| Unvaccinated | 289 (4.2%) |

Table 2 shows the ORs for the association between sociodemographic factors and being unvaccinated among participants who originally intended to vaccinate. In the age-sex-adjusted analysis, compared with participants aged 60–65 years, ORs were significantly higher for those aged 20–29 (OR, 2.62), 30–39 (OR, 3.05), 40–49 (OR, 2.03), and 50–59 years (OR, 1.63). Divorced or widowed participants (OR, 2.72) and singles (OR, 2.43) had significantly higher ORs than married individuals. Regarding annual household income, compared with ≥9 million yen, participants with <4 million (OR, 2.46) and 4–5.99 million yen (OR, 1.70) had significantly higher ORs. Mainly physical workers had a higher OR than mainly desk workers (OR, 1.49). Participants with experience of COVID-19 infection had a higher OR than those without experience (OR, 1.85). In the multivariate-adjusted analysis, participants aged 30–39 (OR, 2.46) and 40–49 years (OR, 1.70), divorced or widowed people (OR, 2.25) and singles (OR, 2.04), participants with an income of <4 million yen (OR 1.63), and those with experience of COVID-19 infection (OR, 2.02) had still significantly higher ORs. We also observed a linear relationship between being unvaccinated and age (*P* for trend = 0.009) and income (*P* for trend = 0.028).

**Table 2.**
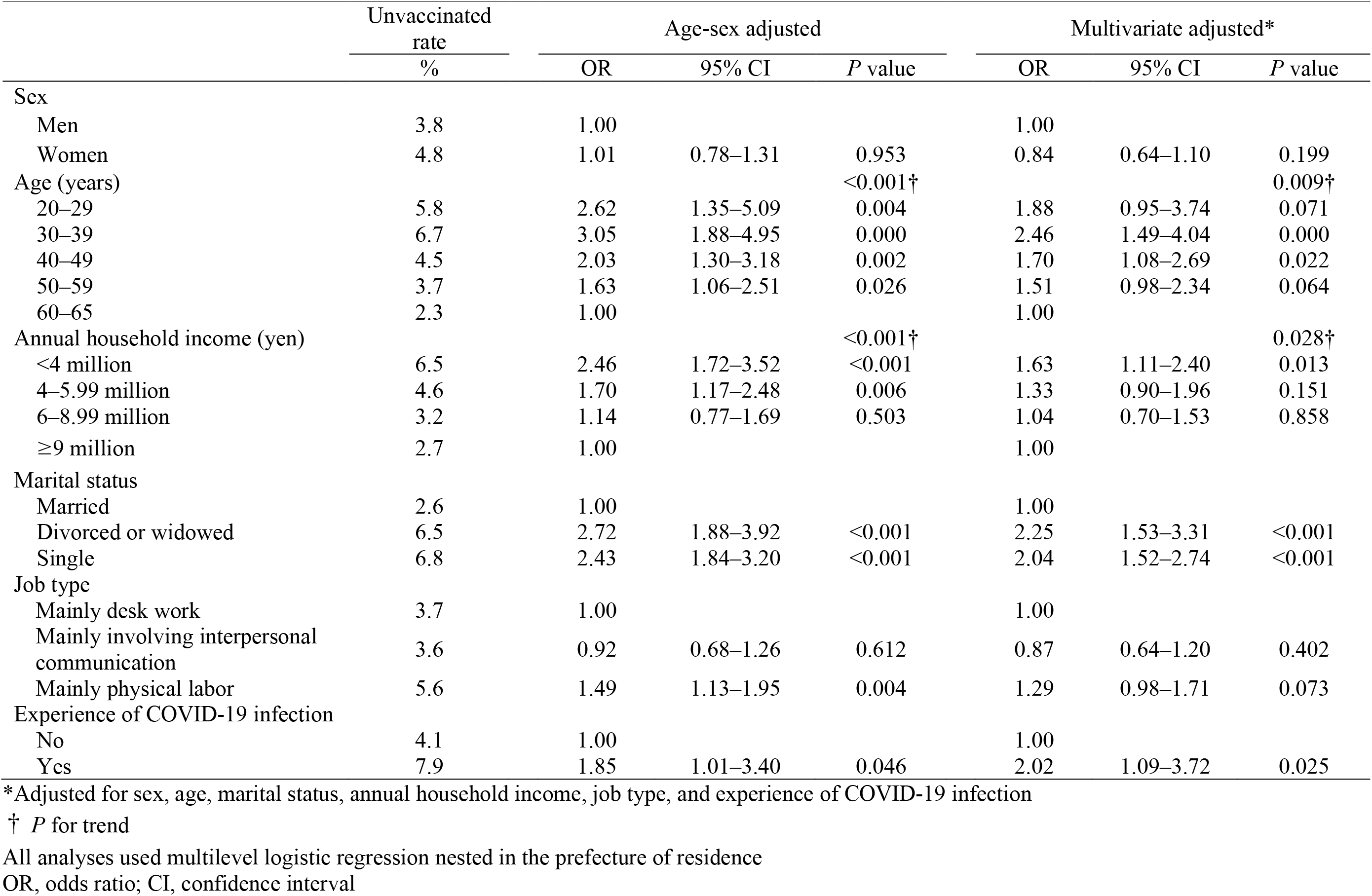
Association between sociodemographic factors and going unvaccinated among participants who originally intended to vaccinate

|  | Unvaccinated<br>rate | Age-sex adjusted |  |  | Multivariate adjusted* |  |  |
| --- | --- | --- | --- | --- | --- | --- | --- |
|  | % | OR | 95% CI | *P* value | OR | 95% CI | *P* value |
| Sex |  |  |  |  |  |  |  |
| Men | 3.8 | 1.00 |  |  | 1.00 |  |  |
| Women | 4.8 | 1.01 | 0.78–1.31 | 0.953 | 0.84 | 0.64–1.10 | 0.199 |
| Age (years) |  |  |  | <0.001† |  |  | 0.009† |
| 20–29 | 5.8 | 2.62 | 1.35–5.09 | 0.004 | 1.88 | 0.95–3.74 | 0.071 |
| 30–39 | 6.7 | 3.05 | 1.88–4.95 | 0.000 | 2.46 | 1.49–4.04 | 0.000 |
| 40–49 | 4.5 | 2.03 | 1.30–3.18 | 0.002 | 1.70 | 1.08–2.69 | 0.022 |
| 50–59 | 3.7 | 1.63 | 1.06–2.51 | 0.026 | 1.51 | 0.98–2.34 | 0.064 |
| 60–65 | 2.3 | 1.00 |  |  | 1.00 |  |  |
| Annual household income (yen) |  |  |  | <0.001† |  |  | 0.028† |
| <4 million | 6.5 | 2.46 | 1.72–3.52 | <0.001 | 1.63 | 1.11–2.40 | 0.013 |
| 4–5.99 million | 4.6 | 1.70 | 1.17–2.48 | 0.006 | 1.33 | 0.90–1.96 | 0.151 |
| 6–8.99 million | 3.2 | 1.14 | 0.77–1.69 | 0.503 | 1.04 | 0.70–1.53 | 0.858 |
| ≥9 million | 2.7 | 1.00 |  |  | 1.00 |  |  |
| Marital status |  |  |  |  |  |  |  |
| Married | 2.6 | 1.00 |  |  | 1.00 |  |  |
| Divorced or widowed | 6.5 | 2.72 | 1.88–3.92 | <0.001 | 2.25 | 1.53–3.31 | <0.001 |
| Single | 6.8 | 2.43 | 1.84–3.20 | <0.001 | 2.04 | 1.52–2.74 | <0.001 |
| Job type |  |  |  |  |  |  |  |
| Mainly desk work | 3.7 | 1.00 |  |  | 1.00 |  |  |
| Mainly involving interpersonal communication | 3.6 | 0.92 | 0.68–1.26 | 0.612 | 0.87 | 0.64–1.20 | 0.402 |
| Mainly physical labor | 5.6 | 1.49 | 1.13–1.95 | 0.004 | 1.29 | 0.98–1.71 | 0.073 |
| Experience of COVID-19 infection |  |  |  |  |  |  |  |
| No | 4.1 | 1.00 |  |  | 1.00 |  |  |
| Yes | 7.9 | 1.85 | 1.01–3.40 | 0.046 | 2.02 | 1.09–3.72 | 0.025 |

\*Adjusted for sex, age, marital status, annual household income, job type, and experience of COVID-19 infection
† *P* for trend
All analyses used multilevel logistic regression nested in the prefecture of residence
OR, odds ratio; CI, confidence interval

## 4. Discussion

We conducted a trajectory study about COVID-19; we examined the sociodemographic factors related to vaccination among participants who at baseline had strong intention to do so. We found that young to middle-aged participants, being divorced, widowed, or single, low-income earners, and individuals with experience of COVID-19 infection were more likely to be unvaccinated. We assumed they became hesitant about vaccination owing to perceived risks associated with vaccine side effects or being unable to receive vaccination for financial reasons.

The reason for becoming hesitant about undergoing vaccination was changing in their risk perceptions related to the vaccines owing to acquiring various information over time. Vaccination decisions are generally influenced by perceptions of two risks: the risk of not being vaccinated (e.g., risk of becoming infected or infection becoming severe); and the risk of vaccination (e.g., vaccine side effects) [14]. COVID-19 vaccines were manufactured shortly after the pandemic was declared; that raised overall concerns about side effects and future safety [15]. Two mRNA vaccines have been used in Japan: one from Pfizer Inc. and one from Moderna Inc. They reportedly caused both local symptoms (such as pain and inflammation at the vaccination site) and systemic symptoms (such as fever, fatigue, and headache) as side effects; those side effects occurred more frequently than with seasonal influenza vaccines, which are administered every year [16], [17], [18], [19]. Thus, we believe those side effects and concerns about vaccine safety owing to the short production time affected the participants’ intentions to vaccinate.

Another factor is that employees may have been obliged to be absent from work owing to vaccination side effects and thus went unvaccinated. In Japan, efforts were made to decrease vaccine hesitancy through access issues by ensuring free vaccination, fair vaccine distribution, and convenient vaccination sites (such as workplace vaccination) [3], [9]. However, compensation for absenteeism on the day of vaccination and through vaccination-related side effects was determined by companies [3], [20]. Therefore, participants who were concerned about vaccination side effects and who were unable or unwilling to take time off work for financial or other reasons may have been less likely to be vaccinated. On the basis of the above two factors, we now discuss each of the study covariates.

We found that younger or middle-aged participants were less likely to be vaccinated than older ones. Younger or middle-aged people were reportedly less likely to be severely infected with COVID-19 than older individuals, which may have reduced infection risk perceptions among the former [21]. The Moderna vaccine had a higher incidence of systemic side effects than the Pfizer vaccine [16]; however, in Japan, the Pfizer vaccine was mainly used in clinics, whereas the Moderna vaccine was applied in mass vaccination by local governments and in occupational vaccination [22]. Accordingly, we observed a higher proportion of young to middle-aged participants who had received the Moderna vaccine—especially among workers, except health-care staff (who were prioritized for vaccination). Further, the suggested incidence of myocarditis and pericarditis in young men owing to these vaccines may have increased concern about side effects [23]. In Japan, it is reported that the Moderna vaccine had a higher incidence than the Pfizer vaccine [22]. Thus, young to middle-aged participants may have received information about COVID-19 infection and vaccines that made them disinclined to undergo vaccination because they believed its risks outweighed the risks of not doing so.

Among low-income earners, we found that many went unvaccinated even though initially they had intended to be vaccinated. Low-income earners have a high unemployment rate; even during COVID-19, sickness presenteeism was reportedly high [24], [25]. It was evidently difficult for such people to take time off work for vaccination. If they did not receive leave compensation for vaccination or its side effects, they may have refused vaccination through concerns about the direct impact on their incomes owing to taking time off work and even about their employment status as a result of absenteeism.

Regarding marital status, we found that single, divorced, or widowed participants were more likely to be unvaccinated than married individuals. This could be due to economic reasons. Japanese married men tend to have higher incomes than unmarried ones [26]. The main causes of divorce in Japan are poor financial circumstances and unstable employment; thus, incomes of divorced or widowed individuals tend to be lower than those of married people [27]. We also observed significant results with the multivariate model. Concerns for the health of family and friends are reportedly linked to vaccination behavior; it is possible that our married participants were more likely to be vaccinated despite the risks associated with vaccination than single, divorced, or widowed people [28].

Regarding job type, we found that manual workers displayed a significant difference in the age-sex model but not in the multivariate model. The results may indicate to have a greater impact on income than work content. Manual workers are mainly occupied on-site, and their production would be affected if they were absent from work. It is also difficult for them to work from home; thus, they would be obliged to be absent from work if vaccination side effects occurred. It has been reported that manual workers show higher sickness presenteeism than teleworkers: it is not easy for the former to take time off work on a daily basis [25]. Manual workers in Japan also have low incomes [29]: it is possible that for them concerns about reduced income due to absenteeism associated with side effects had a major impact on not undergoing vaccination.

With respect to participants with experience of COVID-19 infection going unvaccinated, decreased awareness of the need for vaccination and increased concern about vaccine side effects both possibly had an impact. COVID-19 vaccination is recommended regardless of infection experience, but the reinfection rate reportedly decreases for a while after infection owing to the production of neutralizing antibodies [30], [31]. Further, individuals who have been infected are more likely to have vaccine side effects, which may increase their concerns: they may have slowly acquired this information and chose not to receive vaccination [32], [33]. Others may have missed the opportunity to be vaccinated because they were infected with COVID-19 shortly beforehand—even though they originally planned to be vaccinated.

We found that a small proportion (4%) of respondents did not receive COVID-19 vaccination despite a strong intention to do so. Sociodemographic factors (such as being young to middle-aged; being single, divorced, or widowed; low-income earners; manual workers; and individuals with experience of COVID-19 infection) may have influenced the vaccination decision. Attribute differences in concerns about vaccine side effects and the impact of side effects on work may also have played a role. Our findings offer suggestions to consider when aiming for acquiring herd immunity through vaccination in future epidemics. First, the national government has to consider distributing vaccinations by age according to reports of vaccine side effects and consider the type and location of vaccines being more freely selected. Second, it is necessary to explain repeatedly the benefits and needs for vaccination and maintain or enhance the intention to vaccinate. For individuals with infection experience, it is important to provide appropriate information about the following: antibody production due to infection being low compared with the amount of antibody produced by vaccination; the possibility of reinfection; and vaccination being important even after infection owing to the effects of new mutant strains of SARS-CoV-2 [34], [35]. Third, it is necessary to consider introducing a direct compensation system for workers and a support system for companies so that employees can take leave owing to vaccine side effects.

This study has several limitations. First, we investigated the reasons for participants going unvaccinated despite initially strongly intending to vaccinate; however, the number of people targeted was small, which may have affected the analysis results. Second, the survey was conducted via the Internet, so there is a limit to its generalizability. Notably, some socially vulnerable groups may have been unable to participate because they lacked Internet access for financial reasons. Third, we did not investigate the exact reasons for going unvaccinated: further research in this regard is required. Fourth, we used responses about infection experience up to December 2021, but we also included in our analysis participants with COVID-19 infection after vaccination; thus, our results may be an underestimate.

## 5. Conclusions

We observed that sociodemographic factors (such as being young to middle-aged; being divorced, widowed, or single; having low income; and having COVID-19 infection experience) affect going unvaccinated even if there was initially strong intention to do so. Concerns about vaccine side effects and the impact of side effects on work may have played a role in the vaccination decision. It is necessary to explain repeatedly the need for vaccination and to provide social support to ensure that individuals who intend to vaccinate are able to do so.

## Data Availability

Data not available due to ethical restrictions

## Acknowledgements

We thank the current members of the CORoNaWork Project, in alphabetical order, are as follows: Dr. Akira Ogami, Dr. Ayako Hino, Dr. Hajime Ando, Dr. Hisashi Eguchi, Dr. Keiji Muramatsu, Dr. Koji Mori, Dr. Kosuke Mafune, Dr. Makoto Okawara, Dr. Mami Kuwamura, Dr. Mayumi Tsuji, Dr. Ryutaro Matsugaki, Dr. Seiichiro Tateishi, Dr. Shinya Matsuda, Dr. Tomohiro Ishimaru, and Dr. Tomohisa Nagata, Dr. Yoshihisa Fujino (present chairperson of the study group), and Dr. Yu Igarashi. All members are affiliated with the University of Occupational and Environmental Health, Japan.

## References

[1] Bartsch SM, O’Shea KJ, Ferguson MC, Bottazzi ME, Wedlock PT, Strych U, et al. Vaccine Efficacy Needed for a COVID-19 Coronavirus Vaccine to Prevent or Stop an Epidemic as the Sole Intervention. Am J Prev Med 2020;59:493–503. https://doi.org/10.1016/j.amepre.2020.06.011.

[2] Nicole Lurie, Melanie Saville, Richard Hatchett, Jane Halton. Developing Covid-19 Vaccines at Pandemic Speed. N Engl J Med 2022:1969–73. https://www.nejm.org/doi/full/10.1056/nejmp2005630.

[3] Prime Minister of Japan and His Cabinet. Novel coronavirus vaccines, https://japan.kantei.go.jp/ongoingtopics/vaccine.html [accessed 23 July 2022]

[4] Mahase E. Covid-19: Pfizer vaccine efficacy was 52% after first dose and 95% after second dose, paper shows. BMJ 2020;371:m4826. https://doi.org/10.1136/bmj.m4826.

[5] Dickerman BA, Gerlovin H, Madenci AL, Kurgansky KE, Ferolito BR, Figueroa Muñiz MJ, et al. Comparative Effectiveness of BNT162b2 and mRNA-1273 Vaccines in U.S. Veterans. N Engl J Med 2022;386:105–15. https://doi.org/10.1056/nejmoa2115463.

[6] Falsey AR, Sobieszczyk ME, Hirsch I, Sproule S, Robb ML, Corey L, et al. Phase 3 Safety and Efficacy of AZD1222 (ChAdOx1 nCoV-19) Covid-19 Vaccine. N Engl J Med 2021;385:2348–60. https://doi.org/10.1056/nejmoa2105290.

[7] Fine P, Eames K, Heymann DL. “Herd immunity”: A rough guide. Clin Infect Dis 2011;52:911–6. https://doi.org/10.1093/cid/cir007.

[8] MacDonald NE, Eskola J, Liang X, Chaudhuri M, Dube E, Gellin B, et al. Vaccine hesitancy: Definition, scope and determinants. Vaccine 2015;33:4161–4. https://doi.org/10.1016/j.vaccine.2015.04.036.

[9] Prime Minister of Japan and His Cabinet. Vaccinating Order, https://japan.kantei.go.jp/ongoingtopics/pdf/202105_vaccinating_order.pdf [accessed 23 July 2022]

[10] Digital Agency. Vaccination Record System [in Japanese], https://info.vrs.digital.go.jp/ [accessed 23 July 2022]

[11] Siegler AJ, Luisi N, Hall EW, Bradley H, Sanchez T, Lopman BA, et al. Trajectory of COVID-19 Vaccine Hesitancy over Time and Association of Initial Vaccine Hesitancy with Subsequent Vaccination. JAMA Netw Open 2021;4:5–9. https://doi.org/10.1001/jamanetworkopen.2021.26882.

[12] Fujino Y, Ishimaru T, Eguchi H, Tsuji M, Tateishi S, Ogami A, et al. Protocol for a Nationwide Internet-based Health Survey of Workers During the COVID-19 Pandemic in 2020. J UOEH 2021;43:217–25. https://doi.org/10.7888/JUOEH.43.217.

[13] Organisation for Economic Co-operation and Development, Exchange rates (indicator). https://doi.org/10.1787/037ed317-en [accessed 23 July 2022]

[14] Anderson LR, Mellor JM. Predicting health behaviors with an experimental measure of risk preference. J Health Econ 2008;27:1260–74. https://doi.org/10.1016/j.jhealeco.2008.05.011.

[15] Lin C, Tu P, Beitsch LM. Confidence and receptivity for covid-19 vaccines: A rapid systematic review. Vaccines 2021;9:1–32. https://doi.org/10.3390/vaccines9010016.

[16] Polack FP, Thomas SJ, Kitchin N, Absalon J, Gurtman A, Lockhart S, et al. Safety and Efficacy of the BNT162b2 mRNA Covid-19 Vaccine. N Engl J Med 2020;383:2603–15. https://doi.org/10.1056/nejmoa2034577.

[17] Jackson LA, Anderson EJ, Rouphael NG, Roberts PC, Makhene M, Coler RN, et al. An mRNA Vaccine against SARS-CoV-2 — Preliminary Report. N Engl J Med 2020;383:1920–31. https://doi.org/10.1056/nejmoa2022483.

[18] Anderson EJ, Rouphael NG, Widge AT, Jackson LA, Roberts PC, Makhene M, et al. Safety and Immunogenicity of SARS-CoV-2 mRNA-1273 Vaccine in Older Adults. N Engl J Med 2020;383:2427–38. https://doi.org/10.1056/nejmoa2028436.

[19] Ministry of Health, Labour and Welfare, Japan. Q&A on Influenza-Q.33, https://www.mhlw.go.jp/bunya/kenkou/kekkaku-kansenshou01/qa_eng.html [accessed 23 July 2022]

[20] Ministry of Health, Labour and Welfare, Japan. Q&A about the COVID-19 (for companies) [in Japanese], https://www.mhlw.go.jp/stf/seisakunitsuite/bunya/kenkou_iryou/dengue_fever_qa_00007.html#Q4-20 [accessed 23 July 2022]

[21] Tsuchihashi Y, Arima Y, Takahashi T, Kanou K, Kobayashi Y, Sunagawa T, et al. Clinical Characteristics and Risk Factors for Severe Outcomes of Novel Coronavirus Infection, January–March 2020, Japan, J Epidemiol. 2021; 31(8): 487–494. https://doi.org/10.2188/jea.JE20200519

[22] Ministry of Health, Labour and Welfare, Japan. Evaluation of the Safety of Myocarditis-Related Events Related to the COVID-19 Vaccine [in Japanese], https://www.mhlw.go.jp/content/10601000/000914103.pdf [accessed 23 July 2022]

[23] Klein N, Permanente K. Myocarditis analyses in the Vaccine Safety Datalink : rapid cycle analyses and “head-to-head” product comparisons 2021;202113.

[24] Kuroishi M, Nagata T, Hino A, Tateishi S, Ogami A, Tsuji M, et al. Prospective Cohort Study of Sociodemographic and Work-Related Factors and Subsequent Unemployment under COVID-19 Pandemic. Int. J. Environ. Res. Public Health 2022, 19(11), 6924. https://doi.org/10.3390/ijerph19116924.

[25] Masuda M, Ishimaru T, Hino A, Ando H, Tateishi S, Nagata T, et al. A Cross-Sectional Study of Psychosocial Factors and Sickness Presenteeism in Japanese Workers during the COVID-19 Pandemic. J Occup Environ Med 2022;64:E1–7. https://doi.org/10.1097/JOM.0000000000002415.

[26] Gender Equality Bureau Cabinet Office, Japan. Basic Data on Marriage and Family in Japan [in Japanese], https://www.gender.go.jp/kaigi/kento/Marriage-Family/8th/pdf/1.pdf [accessed 23 July 2022]

[27] Matsuura H, Nakano M, Ito Y. Social Factors ‘Analysis of Japanese Divorce Social Factors’ Analysis of Japanese Divorce n.d.

[28] Turk E, Čelik T, Smrdu M, Šet J, Kuder A, Gregorič M, et al. Adherence to COVID-19 mitigation measures: The role of sociodemographic and personality factors. Curr Psychol 2021. https://doi.org/10.1007/s12144-021-02051-5.

[29] Ministry of Health, Labour and Welfare, Japan. Basic Survey of the Wage Structure in Japan 2020 [in Japanese], https://www.mhlw.go.jp/toukei/itiran/roudou/chingin/kouzou/z2020/dl/13.pdf [accessed 23 July 2022]

[30] Centers for Disease Control and Prevention. Frequently Asked Questions about COVID-19 Vaccination Updated 12 May 2021, https://www.cdc.gov/coronavirus/2019-ncov/vaccines/faq.html [accessed 23 July 2022]

[31] Yamayoshi S, Yasuhara A, Ito M, Akasaka O, Nakamura M, Nakachi I, et al. Antibody titers against SARS-CoV-2 decline, but do not disappear for several months. EClinicalMedicine 2021;32:100734. https://doi.org/10.1016/j.eclinm.2021.100734.

[32] Krammer F, Srivastava K, Alshammary H, Amoako AA, Awawda MH, Beach KF, et al. Antibody Responses in Seropositive Persons after a Single Dose of SARS-CoV-2 mRNA Vaccine. N Engl J Med 2021;384:1372–4. https://doi.org/10.1056/nejmc2101667.

[33] Beatty AL, Peyser ND, Butcher XE, Cocohoba JM, Lin F, Olgin JE, et al. Analysis of COVID-19 Vaccine Type and Adverse Effects Following Vaccination. JAMA Netw Open 2021;4:1–13. https://doi.org/10.1001/jamanetworkopen.2021.40364.

[34] Walls AC, Sprouse KR, Bowen JE, Joshi A, Franko N, Navarro MJ, et al. SARS-CoV-2 breakthrough infections elicit potent, broad, and durable neutralizing antibody responses. Cell 2022;185:872-880.e3. https://doi.org/10.1016/j.cell.2022.01.011.

[35] Centers for Disease Control and Prevention. SARS-CoV-2 Variant Classifications and Definitions, https://www.cdc.gov/coronavirus/2019-ncov/variants/variant-classifications.html [accessed 23 July 2022]

